# Does living in major towns favor institutional delivery in Somalia?

**DOI:** 10.1101/2022.04.23.22274202

**Authors:** Naima Said Sheikh, Ahmed M. Hussein, Shukri Said Mohamed, Abdi Gele

**Affiliations:** Department of Health Services, Norwegian Institute of Public Health, PO Box 222, Skøyen, 0213 Oslo, Norway; Department of Maternal and Reproductive Health, Somali Institute for Health Research, Somalia

## Abstract

**Background:** An institutional delivery is a childbirth that takes place at a health facility in which the birth is assisted by a skilled healthcare provider. Institutional delivery could reduce approximately 33% of maternal deaths. However, the use of institutional healthcare is failing in many Sub-Saharan African countries because of many factors, including poverty, a lack of access, distance, a lack of transport and other socio-cultural factors. In Somalia, only 32% of births are delivered in a health facility with the assistance of a skilled healthcare provider. We aim to investigate the factors hindering women from giving birth at health facilities in major towns in Somalia, where most of the health facilities in the country are concentrated.

**Methods:** A community-based health survey was carried out from 11 major towns in Somalia between October and December 2021. A structured and pretested questionnaire was used to collect data from 430 women who gave birth last five years. A logistic regression analysis was carried out to establish the association between the covariates of interest and the outcome variable.

**Results:** The overall prevalence of institutional delivery was 57%. Approximately 38% of women who live in Mogadishu and 53% of women living in another ten towns give birth at home. Women who had a poor knowledge of the importance of health facility delivery had nearly four times higher odds of delivering at home (OR 3.645 CI: 1.488-8.928). Similarly, those who did not receive antenatal care (OR 2.551, CI: 1.017-6.399), and those who did not receive a consultation on the place of delivery (OR 2.145, CI: 1.167-3.942) were more likely to give birth at home. The reasons for home delivery included financial reasons, must use transport to reach the nearest health facility and it is easier to deliver at home.

**Conclusion:** The study shows that home delivery is high in major towns in Somalia. It is important for health providers to communicate with women and men about the risks related to pregnancy and educate them about the importance of a health facility delivery. Antenatal care should be considered universal for pregnant women, while central and federal governments should guarantee access to free and within-reach ANC for women and girls. In conflict settings in Somalia, this should be done by training community health workers and auxiliary nurses who provide ANC for women through home visits.

## Background

Over half a million women experience pregnancy-related deaths each year, with 99% of these deaths occurring in low- and middle-income countries (LMIC), where complications of pregnancy and childbirth are the leading cause of death among women of reproductive age (1). Approximately 85% of these deaths occur in sub-Saharan Africa and South Asia, with countries experiencing scarce skilled health staff taking the greatest burden (2). The risk of maternal death has declined in some of these countries over the last decade, but there has either been no or diminutive improvement in some countries, including Somalia. More than 70% of all maternal deaths are due to preventable conditions such as hemorrhage, infection, unsafe abortion, hypertensive disorders of pregnancy and obstructed labor (3, 4). To help improve maternal health, adequate access to healthcare during and after pregnancy is necessary (5). However, access to these services is not possible for some women due to underlying causes, such as poverty, inadequate, inaccessible or unaffordable healthcare, unequal access to resources, the low status of women and illiteracy (3).

Armed conflicts are widely held to hinder the progress toward maternal mortality reduction through restricting access to health services and the breakdown of health systems, which can cause a dramatic rise in preventable deaths for mothers during pregnancy and childbirth (6). Recently published literature confirms that mothers who were exposed to armed conflict had a higher likelihood of experiencing maternal deaths (7). Armed conflict is often linked to the lack of- or limited access to maternal healthcare services due to safety, financial and geographical restrictions (8).

Reaching the aim of Sustainable Development Goals (SDG) to reduce maternal and infant mortalities by 2030 (4) requires increased institutional delivery (5). Institutional delivery (ID) could reduce 16% to 33% of maternal deaths through the provision of safe delivery, and reducing complications related to- and occurring during birth (9). Given the verified health benefits of institutional delivery, it is crucial to comprehend the range of factors that determines the choice of the place of childbirth. Prior studies on healthcare use and a subsequent health facility delivery, have highlighted a range of potential influences on a woman’s tendency to seek care at a health facility, which include younger maternal age, a woman and husband’s education, employed, contraception use, previous delivery at a health facility and the utilization of antenatal care (10-12). Despite this knowledge, the use of institutional healthcare delivery is failing in Sub-Saharan Africa due to a lack of access, distance and insufficient equipment (5, 13).

Somalia is a sub-Saharan African country characterized by over three decades of civil war. The maternal mortality rate (MMR) in Somalia is 692 per 100,000, which is one of the highest in the world (14). This is because the risk factors for MMR are abundant in Somalia. The contributing factors for such a high MMR include sociopolitical instability and ongoing wars, as well as other sociocultural challenges, poverty, long distances to the health facility and health system limitations (5). There are also underlying factors, including a high rate of teenage pregnancy, as 20% of girls from 15-19 have already had their first pregnancy, a high fertility rate (6.9 children per women), a low uptake of modern methods of family planning (1%) and a high prevalence of female genital cutting (99%) (15). While the Demographic Health Survey (SDHS) (15) briefly mentioned the level of institutional delivery in Somalia, little is known about the factors related to the utilization of institutional delivery/skilled attendance at delivery in Somalia. The SDHS shows that 32% of births in Somalia (including pastoralists and rural population) are assisted by a skilled healthcare provider (15). To help contribute in filling this gap of knowledge, the aim of this study was to investigate the potential factors associated to institutional delivery among women living in major towns in Somalia, with the ultimate goal of providing relevant policy and programmatic recommendations as we prepare Somalia for achieving the SDG’s health agenda for 2030.

## Methods

### Study design and participants

A community-based survey was carried out in eleven major towns in Somalia between October and December 2021. The study was the first of its kind that investigated the factors associated with institutional delivery in Somalia, which is a key intervention in averting the risk of [maternal mortality](https://www.drishtiias.com/daily-updates/daily-news-analysis/declining-maternal-mortality-ratio-in-india) due to childbirth-related complications. A total of 430 women of childbearing age were interviewed. A pre-tested structured questionnaire was used for the data collection, with trained research assistants with laptops filling out the questionnaires for the women. We included women of reproductive age who gave birth in the last five years, and who live in Mogadishu, Kismayo, Jowhar, Galkayo, Dusomareb, Guriel, Beledwein, Bruao, Hargeisa, Afgoi and Bardere. We used a modified version of a questionnaire that was developed and used for a prior study among Somalis in Ethiopia (16). Initially, we prepared the questionnaire in English, then translated it to the Somali language, and then back translated it to English to ensure the specificity of the questions. The Somali version was pretested with eight women by pretrained research assistants. The necessary adjustment was made to develop the final version. The questionnaires included socio-demographic details, as well as questions that assessed the choice of women regarding where to give birth. The independent variable was selected from the available literature. Our covariate of interest included socio-demographics, such as women’s education, the husband’s education and employment. We also collected variables such as whether women sought antenatal care, postnatal care and child vaccines, in addition to their knowledge about the benefits of institutional delivery, and if they received a consultation about where to give birth.

### Statistical analysis

A statistical analysis was carried out using STATA version 16, and the data were descriptively summarized. As the level of education differed between women and their husbands, we collapsed university and secondary education together for women, while we coded separately for their husbands. We asked if the women knew the importance of the health facility delivery, with the response being coded (yes; 0) and (no; 1). The outcome variable was: Where did you give birth for your last pregnancy?, which was categorized as “health facility” (coded: 0) and “home” (coded: 1). As the data was not normally distributed, Chi-square tests (crosstabs) were employed in calculating group differences and logistic regression to establish the association between the covariates of interest and the outcome variable. We reported the Odds ratio (OR) and 95% confidence intervals (95% CI), with p < 0.05 being considered statistically significant.

### Operational definitions

#### Institutional delivery

This is a delivery assisted by a skilled birth attendant in the health facility.

#### Home delivery

When a mother gave birth at her home or other’s home (neighbor, relatives or family), or when a birth takes place outside the health institution.

## Results

Table 1 shows the summary of descriptive statistics and group differences by place of childbirth. A total of 430 women of childbearing age are included. Approximately 70% of the women reside in Mogadishu, while the rest are living in other towns. Of them, 184 (43%) gave birth at home during their last pregnancy. Approximately 38% of women who live in Mogadishu, and 53% of women living in other towns, gave birth at home. The reasons for home delivery included that “it was easier to deliver at home”; “financial reasons” and a “long distance to the health facility”. A majority of the women (82%) were younger than 35 years old, with approximately 69% being unemployed. Regarding education, 62% (267) of the women and 42% (182) of their husbands had no formal education.

**Table 1:** Group differences by place of delivery.

| Variables |  | Health facility<br>N (%) | Home<br>N (%) | Total<br>N (%) | P-value |
| --- | --- | --- | --- | --- | --- |
| Age of woman in years | 15-25 | 94 (38) | 69 (38) | 163 (38) | 0.482 |
|  | 26-35 | 112 (46) | 77 (42) | 189 (44) |  |
|  | 36 and over | 40 (16) | 38 (21) | 78 (18) |  |
| Civil status | Married | 207 (84) | 155 (84) | 362 (84) | 0.979 |
|  | Divorced or widowed | 39 (16) | 29 (16) | 68 (16) |  |
| Occupation | Employed | 83 (34) | 50 (27) | 133 (31) | 0.145 |
|  | Unemployed | 163 (66) | 134 (73) | 297 (69) |  |
| Women's education | High school and above | 68 (28) | 21 (11) | 89 (21) | 0.000 |
|  | Primary and middle school | 53 (22) | 21 (11) | 74 (17) |  |
|  | Uneducated | 125 (51) | 142 (77) | 267 (62) |  |
| Husband's education | University level | 66 (27) | 15 (8) | 81 (19) | 0.000 |
|  | High school graduate | 40 (16) | 27 (15) | 67 (16) |  |
|  | Primary and middle school | 29 (12) | 17 (9) | 46 (11) |  |
|  | Uneducated | 84 (34) | 98 (53) | 182 (42) |  |
|  | Don't know | 27 (11) | 27 (15) | 54 (13) |  |
| Distance to a health clinic | Walk distance | 134 (54) | 77 (42) | 211 (49) | 0.010 |
|  | Must use transport | 112 (46) | 107 (58) | 219 (51) |  |
| Number of children | 1-3 | 108 (49) | 79 (47) | 187 (84) | 0.762 |
|  | 3-6 | 71 (32) | 60 (36) | 131 (34) |  |
|  | 7 and above | 40 (18) | 28 (17) | 68 (18) |  |
| Sought antenatal care services | Yes | 212 (86) | 67 (36) | 279 (65) | 0.000 |
|  | No | 34 (14) | 117 (64) | 151 (35) |  |
| Received advice about the place of delivery | Yes | 169 (78) | 46 (56) | 215 (72) | 0.000 |
|  | No | 47 (22) | 36 (44) | 83 (28) |  |
| Know the benefits of health facility delivery | yes | 235 (96) | 111 (60) | 346 (81) | 0.000 |
|  | No | 11 (4) | 73 (40) | 84 (20) |  |
| Wants to vaccinate their child | Yes | 236 (96) | 163 (89) | 399 (93) | 0.004 |
|  | No | 10 (4) | 21 (11) | 31 (7) |  |
| Wants to use postnatal care | Yes | 236 (96) | 158 (87) | 394 (92) | 0.001 |
|  | No | 10 (4) | 23 (13) | 33 (8) |  |
| Region of residence (state) | Benadir state (capital city of Somalia) | 184 (75) | 114 (62) | 298 (69) | 0.004 |
|  | Other states | 62 (25) | 70 (38) | 132 (31) |  |

*N: number, %: percentages*

There is a significant difference in education between women who delivered at health facilities, and those who delivered at home (P=0.000). Approximately 77% of women who delivered at home did not receive any formal education. Similarly, 53% of women who delivered at home, and 34% of women who delivered at a health facility, had husbands with no formal education, and this difference was significant (P=0.001). Moreover, 96% of women who knew the benefit of a health facility delivery gave birth at a health facility (P=0.000).

As shown in Table 2, after an adjustment for age, marital status, region of residence and employment, a low level of women’s and their husband’s education had four times and five times higher odds of giving birth at home, respectively. Women who must use transport to access the nearest clinic had an almost two times odds of giving birth at home than those who could reach the facility on foot. Furthermore, those who did not have a good knowledge on the benefits of an institutional delivery had 14 times odds of giving birth at home.

**Table 2:** association between health facility delivery and different covariates.

| Dependent variable: Place of delivery |  | Model 1 |  | Model 2 |  |
| --- | --- | --- | --- | --- | --- |
|  | Variables | Unadjusted OR | CI (95%) | Adjusted OR | CI (95%) |
| Women's Education | **High school and above** | **Ref** |  | **Ref** |  |
|  | Primary and middle school | 1.283 | (0.635 – 2.593) | 1.330 | (0.646-2.738) |
|  | Uneducated | 3.678 | (2.133 – 6.345) | 4.222 | (2.371-7.516) |
| Husband's education | **University level** | **Ref** |  | **Ref** |  |
|  | High school | 2.970 | (1.412 – 6.246) | 2.610 | (1.222-5.577) |
|  | Primary and middle school | 2.579 | (1.136 – 5.858) | 2.709 | (1.168-6.283) |
|  | Uneducated | 5.133 | (2.729 – 9.656) | 5.278 | (2.761-10.091) |
|  | Don't know | 4.400 | (2.029 – 9.540) | 4.315 | (1.944-9.580) |
| Distance to a health clinic | **Walking distance** | **Ref** |  | **Ref** |  |
|  | Must use transport | 1.663 | (1.131 – 2.445) | 1.624 | (1.093-2.413) |
| Number of Children | **1-3 children** | **Ref** |  |  |  |
|  | 3-6 children | 1.155 | (0.737 – 1.811) |  |  |
|  | 7-13 children | 0.957 | (0.545 – 1.681) |  |  |
| Sought antenatal care services | **Yes** | **Ref** |  | **Ref** |  |
|  | No | 10.888 | (6.803 – 17.428) | 11.085 | (6.787-18.105) |
| Received information about the place of delivery | Yes | Ref |  | Ref |  |
|  | No | 2.814 | (1.635 – 4.843) | 2.997 | (1.713-5.243) |
| Know the benefits of health facility delivery | **yes** | **Ref** |  | **Ref** |  |
|  | No | 14.050 | (7.169 – 27.537) | 14.015 | (7.019-27.983) |
| Wants to vaccinate the child | Yes | Ref |  | Ref |  |
|  | No | 3.040 | (1.395 – 6.625) | 2.804 | (1.262-6.234) |
| Wants to use postnatal | Yes | Ref |  | Ref |  |
|  | No | 3.435 | (1.592 – 7.414) | 3.263 | (1.492-7.138) |

*Adjusted: Age, marital status, region of residence and employment*

As shown in table 3, we conducted a further analysis in which women’s and their husband’s level of education was also adjusted. Women who had poor knowledge of the importance of a health facility delivery had almost four times odds of delivering at home (OR 3.645 CI: 1.488-8.928). Similarly, those who did not receive antenatal care (OR 2.551, CI: 1.017-6.399), and those who did not get advice on the place of delivery (OR 2.145, CI: 1.167-3.942) were more likely to deliver at home.

**Table 3:**
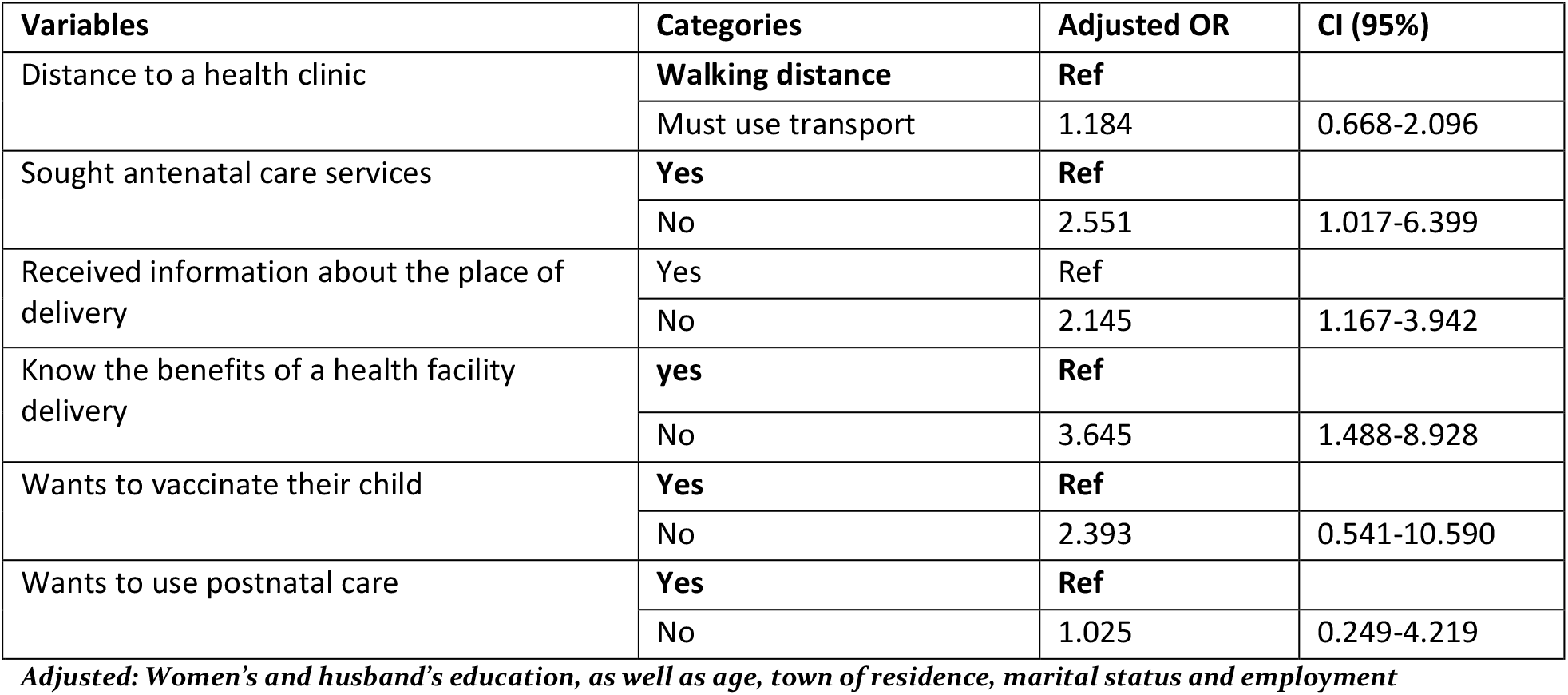
Association between health facility delivery and different covariates after adjustment with education.

| Variables | Categories | Adjusted OR | CI (95%) |
| --- | --- | --- | --- |
| Distance to a health clinic | **Walking distance** | **Ref** |  |
|  | Must use transport | 1.184 | 0.668-2.096 |
| Sought antenatal care services | **Yes** | **Ref** |  |
|  | No | 2.551 | 1.017-6.399 |
| Received information about the place of delivery | Yes | Ref |  |
|  | No | 2.145 | 1.167-3.942 |
| Know the benefits of a health facility delivery | **yes** | **Ref** |  |
|  | No | 3.645 | 1.488-8.928 |
| Wants to vaccinate their child | **Yes** | **Ref** |  |
|  | No | 2.393 | 0.541-10.590 |
| Wants to use postnatal care | **Yes** | **Ref** |  |
|  | No | 1.025 | 0.249-4.219 |

*Adjusted: Women's and husband's education, as well as age, town of residence, marital status and employment*

## Discussion

The study investigated factors associated with institutional delivery among women living in 11 major towns in Somalia. The study shows that 43% of women in major towns in Somalia delivered at home. In line with our findings, SDHS reported that 49% of women in urban areas, 35% percent of women in rural areas and 9% of women in nomadic settlements had received antenatal care (ANC), which is critical for institutional delivery (15). In contrast, the SDHS showed 78% of Somali women had delivered at home in past childbirths. The reason for the higher rates of home delivery in SDHS study is that they did not stratify the variable “place of childbirth” into urban, rural and nomads, which obscured the difference between women who live in major towns, where healthcare facilities are concentrated, and rural and nomadic settlements where access to services are extremely limited. The access to healthcare varies considerably between urban and rural Somalia, as available health services are confined to major towns (17). Our findings are also in agreement with prior studies in Ethiopia, in which the odds of delivering at a health facility was seven times higher for women residing in urban versus rural areas (18).

Women who had no formal education (62%) had over four times higher odds of giving birth at home than women with a secondary or higher level of education. This finding is in line with several studies that reported an association between the education of women and a health facility delivery (10, 11, 19). Our results are also in accordance with the SDHS, which shows 68% of Somali women are illiterate, while only 9% of ever-married women are employed (15). A prior study documented that childbirths of women with no education are less likely to be assisted by skilled personnel (26%) than women with higher education levels (83%) (15). Our results also show that women living with an uneducated husband had five times higher odds of giving birth at home than women with a university/college education. In line with our findings, prior research reported an association between women’s choice of childbirth place and their husband’s level of education (20). A study in Nigeria found that women whose husbands had a tertiary level education had an approximate 61% increased probability of a facility delivery (21). It is not only the education of women that is important in their choice of place of childbirth, but also that educated women had a greater probability of delaying marriage until the age of 18 years and of pregnancy until 20 years (22), as well as low fertility and high contraception use (23).

After controlling for the education level of women and their husbands, women with a poor understanding about the benefits of a health facility delivery were nearly four times higher odds of delivering at home than those with good knowledge. Similarly, those who did not receive consultancy of the place of childbirth were more likely to deliver at home. In line with our findings, a prior study in Zambia found that women who know the danger signs in pregnancy are more likely to deliver in a health facility compared to those without such knowledge (24). Similarly, women in rural Mali who were aware about possible pregnancy complications were more likely to give birth in a health facility (25). Knowledge on the importance of childbirth in a health facility determines the women’s birth preparedness and complication readiness, which is a strategy that has been globally endorsed as an essential component of safe motherhood programs to promote institutional delivery, as well as the timely use of skilled maternal and neonatal care.

Our study shows that the need to use transportation to reach the nearest health facility is associated with home delivery. Distance to health facility is an obstacle to institutional delivery among women in Sub-Saharan Africa (10, 11). In our study, distance to the health facility was measured whether the health facility was in walking distance, or that one must use transport to reach the nearest health facility. A study in Senegal associated a lack of transportation and residence more than five km from a health facility with giving birth at home (26). Moreover, in line with our findings, women in Eritrea who reside within 2 km of a health facility were 14 times more likely to deliver in a health facility than women who have to travel for more than two km to reach the nearest health facility (12). In Somalia, where most of the health facilities are private, and thus more costly (17), pregnant women may not bear the multiple costs associated with seeking institutional delivery, including transport to the facility, and childbirth cost, including standard predelivery and postdelivery expenses such as facility fees and doctor fees, and other costs incurred due to their absence at home and children, which when combined may compel many women in major towns to give birth at home. On the other hand, the frequent armed conflict and rampant bandit roadblocks in Mogadishu, where most of the study participants reside, may discourage women from going to health facilities even within two kms for childbirth, but instead women may feel safe in giving birth at home. Furthermore, women may not consider taking transport to seek an institutional delivery in areas such as Somalia, where childbirth is seen as a normal process, and there is a lack of social and cultural acceptability of facility services (27).

Finally, antenatal care (ANC) and postnatal care utilization, as well as a willingness to vaccinate their child, were all associated with health service delivery in our study. Our findings are in accordance with several studies in Sub-Saharan Africa, thereby showing the importance of ANC utilization in health facility delivery. A literature review demonstrated the role of ANC in influencing health institution delivery, hence suggesting that all the elements of ANC are linked to a higher health facility delivery (11). Likewise, a study in Uganda reported that women who used ANC were more likely to give birth in a healthcare facility than women who do not (28). Additionally, a study conducted in Ethiopia found that ANC follow-up was significantly associated with delivery in a healthcare facility, and as the number of antenatal care visits increased the odds of a facility delivery increased (19). A study among Somalis in Ethiopia also shows that women who attended ANC had a two times higher odds of delivering at a healthcare facility (16). It is widely accepted that ANC promotes the health of pregnant women by reducing the risk of adverse pregnancy outcomes, while encouraging skilled birth attendance and postnatal care, as women who attend ANC are more likely to use these services than non-users (21, 29).

This study has several limitations. It was a cross-sectional study, thus no causal relationship. The study sample was comprised of women who live in major towns where healthcare facilities are concentrated. However, the findings underscore that geographical proximity is not enough for women to seek care from health facilities. We did not measure the distance to the health facility in kilometers, but asked them if they had to use transport to access the facility. This may be a limitation because some people may choose to use transport even if the facility is in walking distance. We did not ask whether women who chose to deliver at home would receive help from a health worker at home or not. However, Somalia where there is an extreme shortage of trained midwives, doctors and nurses, the chance of receiving professional assistance at home is limited. Even though that is the case, giving birth at home in Somalia with the presence of a trained midwife cannot compensate for the benefit of delivering in healthcare facility. The reason for this is that if complications arise during childbirth, it is difficult in Somalia to find an emergency transport to the nearest hospital. Therefore, a planned home delivery may not be a recommendable choice for women giving birth in Somalia.

## Conclusion

The study shows that home delivery is high in major towns in Somalia, and is associated with a lack of understanding on the importance of a health facility delivery, not using ANC and not receiving consultancy about where to give birth. The findings highlight that women in major towns in Somalia do not receive the eight critical reproductive health services, such as antenatal care, delivery, post-natal care, health education, counselling, family planning, treatment and immunization. In this light, it is important for health policymakers and health providers to ensure that women receive access to critical reproductive health services, in addition to communicating with women and men about the risk related to pregnancy, and educate them about the importance of a health facility delivery. Antenatal care should be considered as mandatory for pregnant women, while the government should guarantee access to free and within-reach ANC for women and girls from poor families. This should be done by training community health workers and auxiliary nurses who provide care to pregnant women through home visits. A prior randomized controlled trial found that home visits reduced upstream maternal risks, and improved maternal outcomes (30).

## Data Availability

All data produced in the present study are available upon reasonable request to the authors

## Acknowledgments

We would like to thank our research assistants; Omar Mohamed Abdullahi, Shukri Abdullahi Omar, Ahmed Ali Mohamed, Ifrah Ali Hussein, Lul Abdullahi mohamud, Amina Hassan Hirsi, Hamdi Said Mohamed and Aweys Ahmed Mohamed for collecting the data.

## Data availability statement

The raw data supporting the findings of this paper will be made available on demand by the authors, without undue reservation

## Ethics statement

This study was approved by the Ethical committee at Somali Institute For Health Research. The participants provided oral informed consent to participate in the study.

## Authors’ contributions

AG & NS designed the study, conducted the analysis, interpretation of the findings, and drafted the manuscript. AH and SM contributed data collection and critically revised the manuscript. All authors read and approved the final submitted manuscript.

## Funding

This study did not receive any financial support or grants from any funding agency.

## Conflict of Interest

The authors declare that the research was conducted in the absence of any commercial or financial relationships that could be construed as a potential conflict of interest. The authors declate also that they have no other competing interests.

## Notes

### Competing Interest Statement

The authors have declared no competing interest.

### Funding Statement

This study did not receive any funding

